# Plasma Soluble Tumor Necrosis Factor Receptor Concentrations and Clinical Events after Hospitalization: Findings from ASSESS-AKI and ARID studies

**DOI:** 10.1101/2021.11.08.21266015

**Authors:** Steven G. Coca, George Vasquez-Rios, Sherry G. Mansour, Dennis G. Moledina, Heather Thiessen-Philbrook, Mark M. Wurfel, Jonathan Himmelfarb, Eddie Siew, Amit X. Garg, Chi-yuan Hsu, Kathleen D. Liu, Paul L. Kimmel, Vernon M. Chinchilli, James S. Kaufman, Michelle Wilson, Rosamonde E Banks, Rebecca Packington, Eibhlin McCole, Mary Jo Kurth, Ciaran Richardson, Alan S. Go, Nicholas M Selby, Chirag R. Parikh

**Author notes:** **Corresponding Authors:** Steven G. Coca, DO, MS, Nephrology Division, Icahn School of Medicine at Mount Sinai, One Gustave L. Levy Place, Box 1243, New York, NY 10029, or Chirag R. Parikh, MD, PhD, Director, Division of Nephrology, Ronald Peterson Professor of Medicine, Johns Hopkins School of Medicine, 1830 E. Monument St., Suite 416, Baltimore, MD 21287. Co-primary first authors (Steven G. Coca, George Vasquez-Rios).

## Abstract

**Background:** The role of plasma soluble tumor necrosis factor receptor (sTNFR)1 and sTNFR2 in the prognosis of clinical events after hospitalization with or without acute kidney injury (AKI) is unknown.

**Methods:** We measured sTNFR1 and sTNFR2 obtained 3 months post-discharge using samples from Assessment, Serial Evaluation, and Subsequent Sequelae of Acute Kidney Injury (ASSESS-AKI) and AKI Risk in Derby (ARID) that enrolled patients with and without AKI. The associations between biomarkers with longitudinal kidney disease incidence and progression, heart failure and death were evaluated. Analyses were adjusted for demographics and key covariates at the 3-month visit.

**Results:** Among 1474 participants with plasma biomarker measurements, 19% developed kidney disease progression, 14% had later heart failure, and 21% died over a median follow-up of 4.4 years. For the kidney outcome, the adjusted HRs per doubling in concentration were 2.9 (2.2-3.9) for sTNFR1 and 1.9 (1.5-2.5) for sTNFR2. AKI during the index hospitalization did not modify the association between biomarkers and kidney events. For heart failure, the adjusted HRs per doubling in concentration were 1.9 (1.4-2.5) for sTNFR1 and 1.5 (1.2-2.0) for sTNFR2. For mortality, the adjusted HRs were 3.3 (2.5-4.3) for sTNFR1 and 2.5 (2.0-3.1) for sTNFR2. The findings in ARID were qualitatively similar for the magnitude of association between biomarkers and outcomes.

**Conclusion:** Plasma sTNFR1 and sTNFR2 measured 3 months after discharge were independently associated with clinical events, regardless of AKI status during the index admission. sTNFR1 and sTNFR2 may assist with the risk stratification of patients during follow-up.

**Significance Statement:** Soluble tumor necrosis factor receptor 1 (sTNFR1) and sTNFR2 associate with kidney outcomes in patients with chronic kidney disease with and without diabetes mellitus. However, their role in the post-hospitalization stage is unknown. High sTNFR1 and sTNFR2 obtained 3 months after discharge associate with kidney events, heart failure hospitalizations, and death among patients who did and did not have acute kidney injury (AKI). Furthermore, sTNFRs provide discriminative value at the time of predicting kidney events. These findings were demonstrated in two large independent prospective cohorts. sTNFR1 and sTNFR2 may detect patients at risk of future adverse events even when patients do not meet the clinical criteria for AKI or exhibit biochemical abnormalities.

## Introduction

Prediction of kidney outcomes after an acute illness requiring hospitalization is a major challenge in clinical practice. Acute kidney injury (AKI) is prevalent in the hospital and has been recognized as a major contributor to the burden of chronic kidney disease (CKD) in several studies.^1-4^ Yet, it is estimated that only 19% of those who survive a hospitalization involving AKI receive appropriate follow-up within 12 months after discharge.^5-7^ Furthermore, both AKI and CKD independently and concomitantly increase the risk for future cardiovascular complications and death, which intensifies with further kidney function decline over time.^8,9^ These observations support the pursuit to identify biomarkers and clinical strategies to identify patients at the greatest risk of kidney disease progression.

Recently, the Acute Disease Quality Imitative (ADQI) Consensus conference proposed the implementation of damage and functional biomarkers of AKI for prediction and staging of CKD during post-AKI care.^10^ There is growing literature on the prognostic role of soluble tumor necrosis factor receptors (sTNFRs), particularly in kidney-related outcomes.^11-16^ Tumor necrosis factor alpha (TNF-α) signaling pathways play a major role in cellular stress and inflammatory responses through soluble TNFR1 (sTNFR1) and soluble TNFR2 (sTNFR2).^10-11^ High sTNFR1 and sTNFR2 concentrations measured in the hospital have been associated with protracted kidney recovery in critically ill patients within 7-60 days.^17^ However, previous studies have shown that complications associated with AKI can extend beyond 60 days, in parallel to the debilitating effects of an acute hospital admission.^2-4^ Moreover, depending on the etiology, up to 40% of the patients with AKI during hospitalization do not recover kidney function to their previous baseline level.^18,19^ Although the transition to CKD or end-stage kidney disease (ESKD) is typically determined by the 90-day mark, renal recovery has been documented beyond this time frame.^20-24^ Therefore, a number of patients could be left under-monitored and eventually suffer from the burden of CKD and ESKD without the appropriate tools for assessing the varying risk of future clinical events.^23,25^

The Assessment, Serial Evaluation, and Subsequent Sequelae of Acute Kidney Injury (ASSESS-AKI) study collected blood and urine samples at each annual visit to examine the added value of novel biomarkers in the risk-stratification of kidney and cardiovascular outcomes.^26^ The AKI Risk in Derby (ARID) study was an independent prospective UK study that also aimed to study long-term outcomes after AKI in a protocolized fashion.^27^ While the value of measuring plasma sTNFR1 and sTNFR2 to identify patients at risk of kidney function decline and death has been investigated in CKD and diabetes mellitus, their role in the post-hospitalization setting and AKI has not been rigorously evaluated.^14,15^ We sought to further explore the association between plasma sTNFR1 and sTNFR2 concentrations post-discharge with long-term kidney-related outcomes, heart failure hospitalizations, and mortality in ASSESS-AKI and ARID among patients who did and did not have AKI. Furthermore, we aimed to assess whether measuring sTNFR1 and sTNFR2 could add to the performance of clinical models.

## METHODS

### Cohort Descriptions

The primary analyses were conducted in the ASSESS-AKI study; a multicenter, prospective, parallel, matched cohort study. A detailed description of the study design and methods has been published elsewhere.^26^ Briefly, ASSESS-AKI included 1538 hospitalized adult participants that developed AKI and matched individuals who did not have AKI and who survived to complete an in-person visit at 3 months after discharge. Patients in the medical and surgical floors, as well as in the intensive care units who met the study criteria were enrolled. Participants were age ≥18 years and had pre-admission serum creatinine levels that were obtained within 1 year of the index hospitalization in the outpatient setting or non-emergency department visit. Detailed inclusion criteria have been described in the study protocol.^26^ Exclusion criteria included the presence of acute glomerulonephritis, hepatorenal syndrome, multiple myeloma, malignancy, urinary obstruction, severe heart failure, kidney replacement therapy (dialysis, transplant) prior to hospitalization, pregnancy, or predicted survival ≤ 12 months.

AKI during the index hospitalization was defined as a relative increase of ≥50% or absolute increase of ≥0.3 mg/dL in peak inpatient serum creatinine concentration above the baseline creatinine. AKI stage was classified following the Acute Kidney Injury Network (AKIN) guidelines. In parallel, a matched sample of 769 hospitalized adults without AKI were enrolled at each participating site. Absence of AKI was defined as having both <20% relative increase and ≤0.2 mg/dL absolute increase in peak inpatient serum creatinine concentration compared to baseline. Patients were matched in each individual research center by pre-admission CKD status/eGFR (estimated glomerular filtration rate) by using the Chronic Kidney Disease Epidemiology Collaboration (CKD-EPI) equation, demographics, and comorbidities.^28^ Participants enrolled in this study had study visits at 3 and 12 months after discharge, and annually thereafter, with interim phone contacts at 6 month intervals.

The AKI Risk in Derby (ARID) study was an independent prospective cohort investigating the association between AKI among hospitalized patients with long-term outcomes; similar in design to ASSESS-AKI. ARID recruited two subgroups of patients who had been hospitalized at the Royal Derby Hospital (UK) and who survived at least 90 days after hospitalization. Individuals with and without AKI were identified through automated screening of serum creatinine results.^29^ The presence of AKI was identified by Kidney Disease Improving Global Outcomes (KDIGO) criteria. Urine output was not used due to its inaccurate recording in a general hospitalized population. The etiology of AKI was determined by chart review. Biochemistry results of participants in the non-exposed group were reviewed to ensure that they had not sustained AKI during hospitalization. Participants were followed in a protocolized fashion at 3 months, 12 months and 3 years.^30^ ARID clinical endpoints at 3 years were used to compare biomarkers and the risk of CKD incidence/progression, heart failure, and mortality among subgroups at each time point.^27,30^ Outcome data were available for participants with a median (IQR) follow-up period of 4.4 (2.5, 5.7) years in ASSESS-AKI by the time of analysis, while in ARID, all the participants included in the analysis had outcome data collected after a 3-year follow-up. ARID used the last serum creatinine available in records If a patient expired before the 3-year mark.

### Covariates

Demographic characteristics were recorded for each participant. To align covariates, we collapsed race into 2 categories: ‘white’ and ‘non-white’ and included baseline CKD status (yes/no). All clinical variables were obtained from medical records. Biomarker quartiles were defined within each cohort through different diagnostic assays. At baseline visit, occurring within 90 days after discharge, serum creatinine was measured using an isotope dilution mass spectrometry-traceable enzymatic assay (Roche Di-agnostics, Indianapolis, IN), and a random spot urine protein-to-creatinine ratio using a turbidimetric method (Roche) in ASSESS-AKI. Serum creatinine and urinary albumin were measured in ARID via an enzymatic assay and an immunoturbidimetric assay (Tina-quant Albumin Generation 2), both on the Roche Cobas 702 module (Roche Diagnostics Limited, Burgess Hill, West Sussex, UK), respectively. Urinary creatinine was measured by the Jaffe method.

### Biomarker Measurements

In ASSESS-AKI, plasma samples collected at the 3-month post-hospitalization visit underwent a single controlled thaw, were centrifuged at 5000xg for 10 minutes at 4*C, separated into 1 mL aliquots, and immediately stored at -80°C until biomarkers were measured. Plasma sTNFR1 and sTNFR2 were measured in a single batch using the Meso Scale Discovery platform (Meso Scale Diagnostics, Gaithersburg, MD), which uses electrochemiluminescence detection combined with patterned arrays. The intra-assay coefficient of variation (CV) for sTNFR1 was 5.3% with a detection range of 12.70-120,000 pg/mL. The intra-assay CV for sTNFR2 was 9.3%, with a detection range of 3.2-120,000 pg/mL. All laboratory personnel were blinded to the disease status and outcomes of interests.

In ARID, plasma sTNFR1 and sTNFR2 were measured using Randox’s multiplexed Biochip Arrays with an Evidence Investigator™ analyzer according to manufacturer’s instructions (Randox Laboratories Ltd, Crumlin, UK). The biochips use chemiluminescence detection from multiplexed arrays. CKD Biochip Array I was used for measuring sTNFR1 and sTNFR2, alongside other biomarkers. The intra-assay coefficient of variation (CV) for sTNFR1 was 9.08% with a detection range of 0.04-10 ng/mL. The intra-assay CV for sTNFR2 was 5.96%, with a detection range of 0.07-20 ng/mL. No bridging experiments were conducted between platforms.

### Outcome Definitions

The outcomes for the analyses included kidney disease progression, heart failure hospitalization, and death. Kidney disease incidence/progression was defined as a composite of i) incident CKD (no evidence of pre-existing CKD at the index hospitalization and >25% reduction in the eGFR from baseline; or reaching CKD 3 or worse during follow-up) or ii) progression of pre-existing CKD (characterized by eGFR <60 ml/min/1.73m^2^ during hospitalization and >50% reduction in the baseline eGFR; or progression to CKD 5) or iii) development of ESKD on 3-month follow-up (hemodialysis or peritoneal dialysis requirement at least once/week for >12 weeks, receiving a kidney transplant and/or death while on dialysis). Heart failure hospitalization was identified by discharge codes and the Framingham Heart Study criteria in ASSESS-AKI.^31^ In ARID, HF hospitalization was defined by ICD10 discharge codes.

Death was ascertained from proxy reports, medical records, and death certificate data. In the primary analysis, ASSESS-AKI evaluated time to clinical events within a median (IQR) follow-up of 4.4 (IQR 2.5, 5.7), whereas ARID determined the outcomes of kidney disease progression and death at annual intervals. We examined the binary outcomes of the composite of kidney disease progression and death after 3 years of follow-up for each participant in ASSESS-AKI and ARID during the secondary analysis (**Supplemental Figure 1**).

### Statistical analysis

Descriptive statistics for continuous variables were reported as mean (standard deviation) or median [interquartile range (IQR)] when appropriate. Categorical variables were presented as frequencies. Clinical and demographic differences were evaluated through the Kruskal-Wallis test or Chi-square tests. The primary analysis consisted in the evaluation of clinical outcomes including kidney disease progression, heart failure hospitalizations, and mortality in ASSESS-AKI. We used Cox proportional hazard models that were adjusted for race, sex, age, body mass index (BMI), smoking status, history of chronic obstructive pulmonary disease (COPD), cardiovascular disease (CVD), AKI status (vs. no AKI), CKD status (vs. no CKD), sepsis, 3-month eGFR, 3-month urine albumin creatinine ratio (UACR), 3-month c-reactive protein (CRP), and diabetes mellitus status. Hazard ratios (HR) and 95% confidence intervals (CI) multivariable models are reported. In addition, we assessed the receiver operating characteristic (ROC) curves of sTNFR1 and sTNFR2 in ASSESS-AKI for the discrimination of kidney events alone and when added to a clinical model comprised of eGFR, UACR, and C-reactive protein at 3 months, AKI at the index hospitalization, race/ethnicity, age, smoking, and COPD.

In the secondary analysis, we examined the binary outcomes of kidney disease progression, heart failure hospitalizations and mortality through 3 years of follow-up among participants in both ASSESS-AKI and ARID. Logistic regression models were also adjusted for the key covariates described above. Two-tailed P values of less than 0.05 were considered to indicate statistical significance. The analyses were conducted using SAS software, version 9.4 (Cary, NC). This study was approved by the institutional review boards of the participating institutions.

## RESULTS

### Baseline characteristics of the participants in ASSESS-AKI

Of the 1538 participants in ASSESS-AKI, 1474 had plasma sTNFR1 and sTNFR2 measures (**Supplemental Figure 1)**. Among 1474 patients included in the analysis, the mean age was 65 ± 13 years, 63% were male and 82% were white. Patients were followed for a median of 4.4 (IQR 2.5, 5.7) years. Clinical characteristics and major comorbidities by quartiles of plasma sTNFR1 and sTNFR2 are shown in **Table 1** and **Supplementary Table 1**, respectively. At baseline, older age, AKI, and pre-hospitalization CKD were statistically associated with higher biomarker concentrations.

**Table 1.**
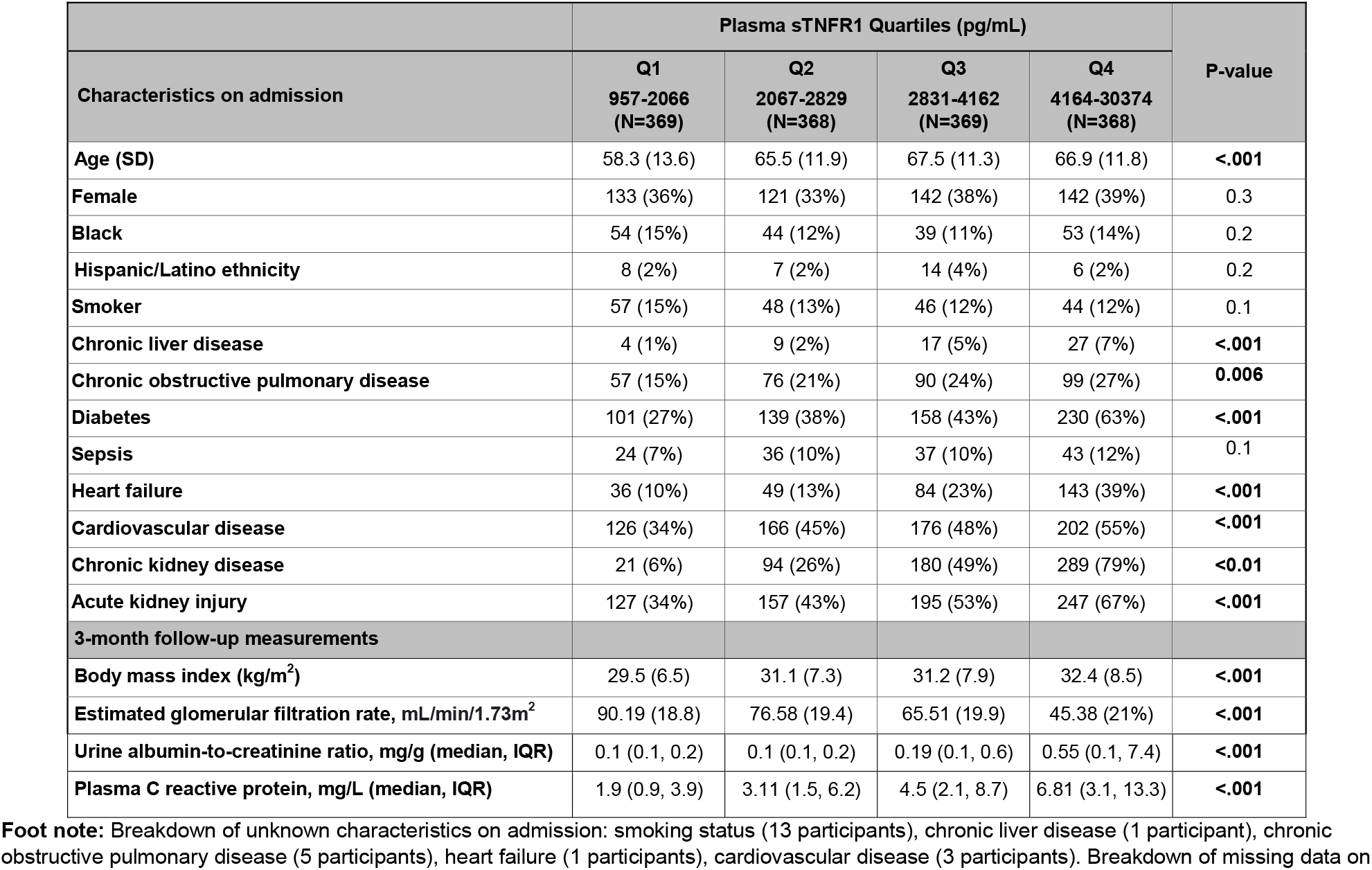
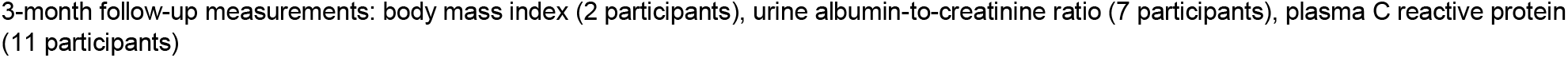
Patients’ characteristics on admission and by 3-month stratified by quartiles of plasma sTNFR1 concentrations in ASSESS-AKI.

|  | Plasma sTNFR1 Quartiles (pg/mL) |  |  |  | P-value |
| --- | --- | --- | --- | --- | --- |
| Characteristics on admission | Q1<br>957-2066<br>(N=369) | Q2<br>2067-2829<br>(N=368) | Q3<br>2831-4162<br>(N=369) | Q4<br>4164-30374<br>(N=368) |  |
| Age (SD) | 58.3 (13.6) | 65.5 (11.9) | 67.5 (11.3) | 66.9 (11.8) | <.001 |
| Female | 133 (36%) | 121 (33%) | 142 (38%) | 142 (39%) | 0.3 |
| Black | 54 (15%) | 44 (12%) | 39 (11%) | 53 (14%) | 0.2 |
| Hispanic/Latino ethnicity | 8 (2%) | 7 (2%) | 14 (4%) | 6 (2%) | 0.2 |
| Smoker | 57 (15%) | 48 (13%) | 46 (12%) | 44 (12%) | 0.1 |
| Chronic liver disease | 4 (1%) | 9 (2%) | 17 (5%) | 27 (7%) | <.001 |
| Chronic obstructive pulmonary disease | 57 (15%) | 76 (21%) | 90 (24%) | 99 (27%) | 0.006 |
| Diabetes | 101 (27%) | 139 (38%) | 158 (43%) | 230 (63%) | <.001 |
| Sepsis | 24 (7%) | 36 (10%) | 37 (10%) | 43 (12%) | 0.1 |
| Heart failure | 36 (10%) | 49 (13%) | 84 (23%) | 143 (39%) | <.001 |
| Cardiovascular disease | 126 (34%) | 166 (45%) | 176 (48%) | 202 (55%) | <.001 |
| Chronic kidney disease | 21 (6%) | 94 (26%) | 180 (49%) | 289 (79%) | <0.01 |
| Acute kidney injury | 127 (34%) | 157 (43%) | 195 (53%) | 247 (67%) | <.001 |
| 3-month follow-up measurements |  |  |  |  |  |
| Body mass index (kg/m<sup>2</sup>) | 29.5 (6.5) | 31.1 (7.3) | 31.2 (7.9) | 32.4 (8.5) | <.001 |
| Estimated glomerular filtration rate, mL/min/1.73m<sup>2</sup> | 90.19 (18.8) | 76.58 (19.4) | 65.51 (19.9) | 45.38 (21%) | <.001 |
| Urine albumin-to-creatinine ratio, mg/g (median, IQR) | 0.1 (0.1, 0.2) | 0.1 (0.1, 0.2) | 0.19 (0.1, 0.6) | 0.55 (0.1, 7.4) | <.001 |
| Plasma C reactive protein, mg/L (median, IQR) | 1.9 (0.9, 3.9) | 3.11 (1.5, 6.2) | 4.5 (2.1, 8.7) | 6.81 (3.1, 13.3) | <.001 |

**Foot note:** Breakdown of unknown characteristics on admission: smoking status (13 participants), chronic liver disease (1 participant), chronic obstructive pulmonary disease (5 participants), heart failure (1 participants), cardiovascular disease (3 participants). Breakdown of missing data on
3-month follow-up measurements: body mass index (2 participants), urine albumin-to-creatinine ratio (7 participants), plasma C reactive protein (11 participants)

### Association between plasma biomarkers and kidney events in ASSESS-AKI and biomarker prognostic performance

Plasma sTNFR1 and sTNFR2 were moderately correlated with eGFR and weakly correlated with CRP and UACR (**Supplementary Table 2**). Among 1474 participants, 19% experienced kidney disease progression, with 13% having incident CKD and 6% progression of pre-existing CKD within a median of 4.4 years (IQR 2.5, 5.7). Fifty participants (3%) reached ESKD after having incident CKD or progression of pre-existing CKD during study follow-up (**Figure 1)**.

**Figure 1.**
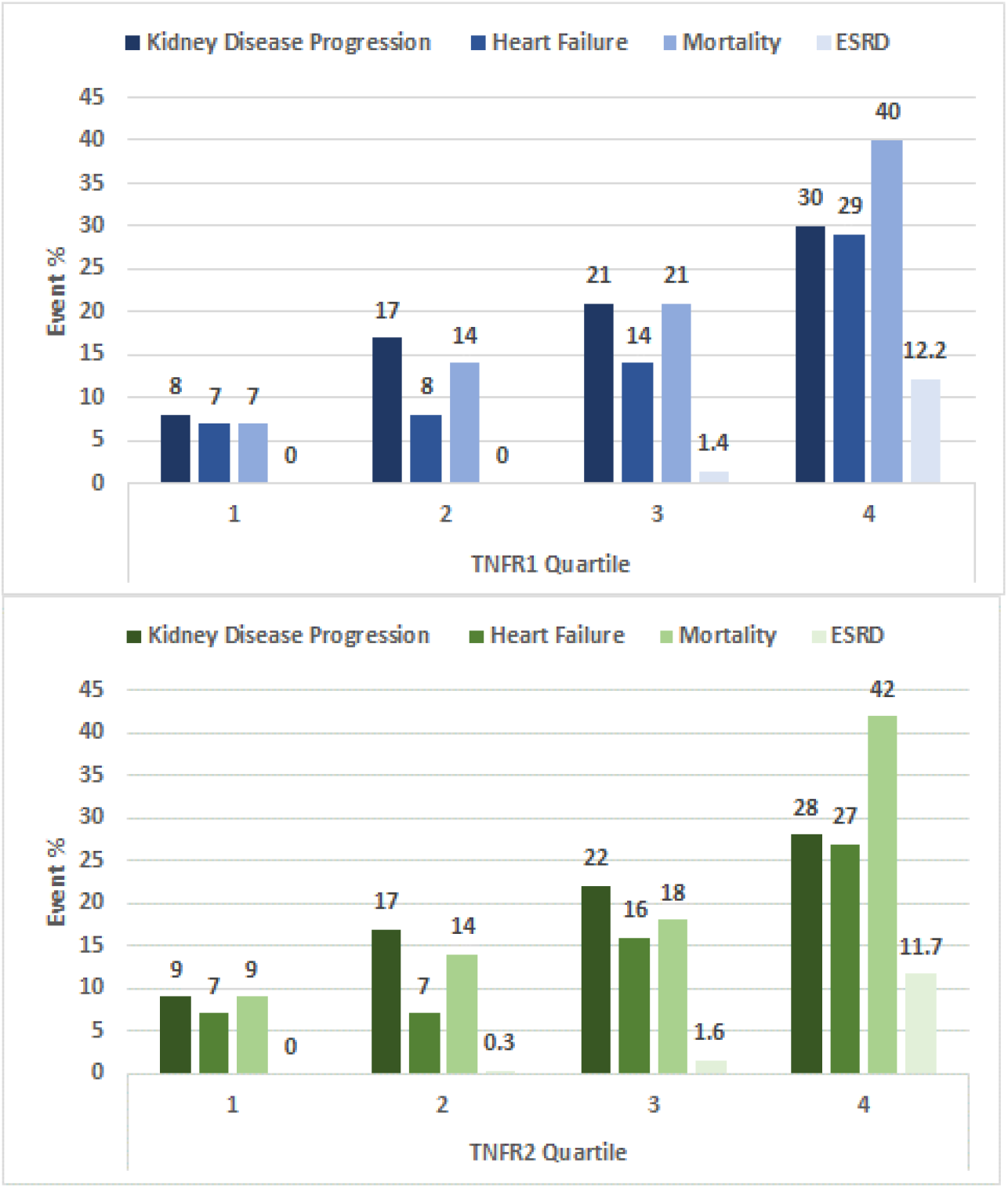
Cumulative incidence of outcomes by quartiles of plasma sTNFR1 and sTNFR2 concentrations in ASSESS-AKI. **Footnote:** 1474 ASSESS-AKI participants followed over a median follow-up of 4.4 (IQR 2.5, 5.7) years.

In adjusted models, plasma biomarkers measured 3 months after discharge were associated with kidney disease progression, independent of AKI status during the index hospitalization and key covariates (**Table 2** and **Figure 2**). The strength of association between kidney disease progression and plasma biomarkers increased linearly. Participants in the highest (4^th^) quartile vs. the lowest (1^st^) quartile of sTNFR1 had nearly 5-fold increased risk of the composite kidney endpoint (adjusted HR: 4.7, 95% CI: 2.6-8.6). Likewise, participants in the 4^th^ (highest) quartile vs. the 1^st^ (lowest) quartile of sTNFR2 were at > 3-fold increased risk of the kidney endpoint (aHR 3.3, 95% CI: 1.9-5.7). Although the absolute incidence of kidney events was relatively higher in those that had AKI compared to those without it, there was no evidence that AKI interacted with the association between biomarkers and the kidney outcome (**Supplemental Table 3**). Furthermore, each doubling in the concentration of sTNFR1 and sTNFR2 was associated with a 11-18-fold higher incidence of ESRD (**Table 2 and Figure 2**). sTNFR1 and sTNFR2 alone at 3 months post-hospitalization had an AUROC of 0.89 (0.85-0.94) and 0.86 (0.81-0.91), respectively, for the composite kidney outcome. AUROC improved to 0.94 (0.93-0.97) and 0.94 (0.91-0.97) for sTNFR1 and sTNFR2, respectively when they were added to basic set of clinical predictors.

**Table 2.** Associations between plasma sTNFR1 and sTNFR2 concentrations with outcomes in ASSESS-AKI.

| Biomarker | Range, pg/mL<br>(Min, Max) | Kidney Disease Progression† |  | Heart Failure |  | All-Cause Mortality |  | ESRD |  |
| --- | --- | --- | --- | --- | --- | --- | --- | --- | --- |
|  |  | Mean (95%CI)<br>event rate per<br>1000 PY | Adjusted*<br>Hazard Ratio<br>(95%CI) | Mean (95%CI)<br>event rate per<br>1000 PY | Adjusted*<br>Hazard Ratio<br>(95%CI) | Mean (95%CI)<br>event rate per<br>1000 PY | Adjusted*<br>Hazard Ratio<br>(95%CI) | Mean (95%CI)<br>event rate per<br>1000 PY | Unadjusted<br>Hazard Ratio<br>(95%CI) |
| sTNFR1<br>(pg/mL) | Log<sub>2</sub><br>Transformed | 46.1 (41, 51.8) | 2.9 (2.2, 3.9) | 33.3 (29, 38.1) | 1.9 (1.4, 2.5) | 45.4 (40.6, 50.8) | 3.3 (2.5, 4.3) | 7.6 (4.7, 10.0) | 18.2 (11.1, 29.80) |
|  | Q1<br>(957, 2066) | 15.9 (11.1, 22.9) | 1 (ref) | 13.3 (8.9, 19.8) | 1 (ref) | 13.9 (9.5, 20.4) | 1 (ref) | 0 |  |
|  | Q2<br>(2067, 2829) | 39.2 (30.7, 50.1) | 2.4 (1.5, 3.9) | 18.1 (12.7, 25.7) | 0.9 (0.6, 1.7) | 29.7 (22.7, 38.9) | 1.9 (1.1, 3) | 0 |  |
|  | Q3<br>(2831, 4162) | 53.5 (42.9, 66.8) | 3.8 (2.3, 6.4) | 31.8 (24.1, 41.9) | 1.3 (0.8, 2.3) | 46.7 (37.4, 58.3) | 2.6 (1.5, 4.2) | 3.0 (1.3, 7.2) |  |
|  | Q4<br>(4164, 30374) | 93.1 (77.1, 112.3) | 4.7 (2.6, 8.6) | 86.3 (71.3, 104.5) | 2.3 (1.3, 4.3) | 104.9 (89.3, 123.2) | 4.9 (2.8, 8.6) | 34.4 (25.7, 46.1) |  |
| sTNFR2<br>(pg/mL) | Log<sub>2</sub><br>Transformed | 46.1 (41, 51.8) | 1.9 (1.5, 2.5) | 33.3 (29, 38.1) | 1.5 (1.2, 2) | 45.4 (40.6, 50.8) | 2.5 (1.9, 3.1) | 7.6 (4.7, 10.0) | 10.9 (7.07, 17.08) |
|  | Q1<br>(2022, 4756) | 18.4 (13.1, 25.9) | 1 (ref) | 14.6 (9.9, 21.4) | 1 (ref) | 17.4 (12.3, 24.6) | 1 (ref) | 0 |  |
|  | Q2<br>(4758, 6651) | 39.1 (30.4, 50.1) | 1.9 (1.2, 3.2) | 15.3 (10.4, 22.4) | 0.8 (0.4, 1.3) | 29.7 (22.6, 39) | 1.5 (0.9, 2.4) | 0.6 (0.1, 4.1) |  |
|  | Q3<br>(6652, 9693) | 54.5 (43.8, 67.8) | 3.3 (2.1, 5.2) | 38 (29.5, 49) | 1.4 (0.8, 2.3) | 39.5 (31.1, 50.2) | 1.7 (1.1, 2.7) | 3.6 (1.6, 7.9) |  |
|  | 4<sup>th</sup><br>(9710, 53522) | 85.8 (70.8, 104) | 3.3 (1.9, 5.7) | 78.2 (64.1, 95.3) | 1.9 (1.1, 3.3) | 106.8 (91.2, 125) | 3.9 (2.4, 6.3) | 32.0 (23.7, 43.2) |  |

**Footnote:** Analysis obtained within a median of 4.4 years (IQR2.5, 5.7).
CI: confidence interval; PY: person-year; sTNFR1: soluble tumor necrosis factor α receptor 1; sTNFR2: soluble tumor necrosis α receptor 2.
†Kidney disease progression is the composite outcome including chronic kidney disease incidence, progression of pre-existing chronic kidney disease and end-stage kidney disease.
\*Adjusted for race, sex, age, body mass index at 3 months, smoking status, history COPD, CVD, AKI status (vs. no AKI), CKD status (vs. no CKD), sepsis during hospitalization, 3-month eGFR, 3-month UACR, C-Reactive protein at 3 months, and clinical site

**Figure 2.**
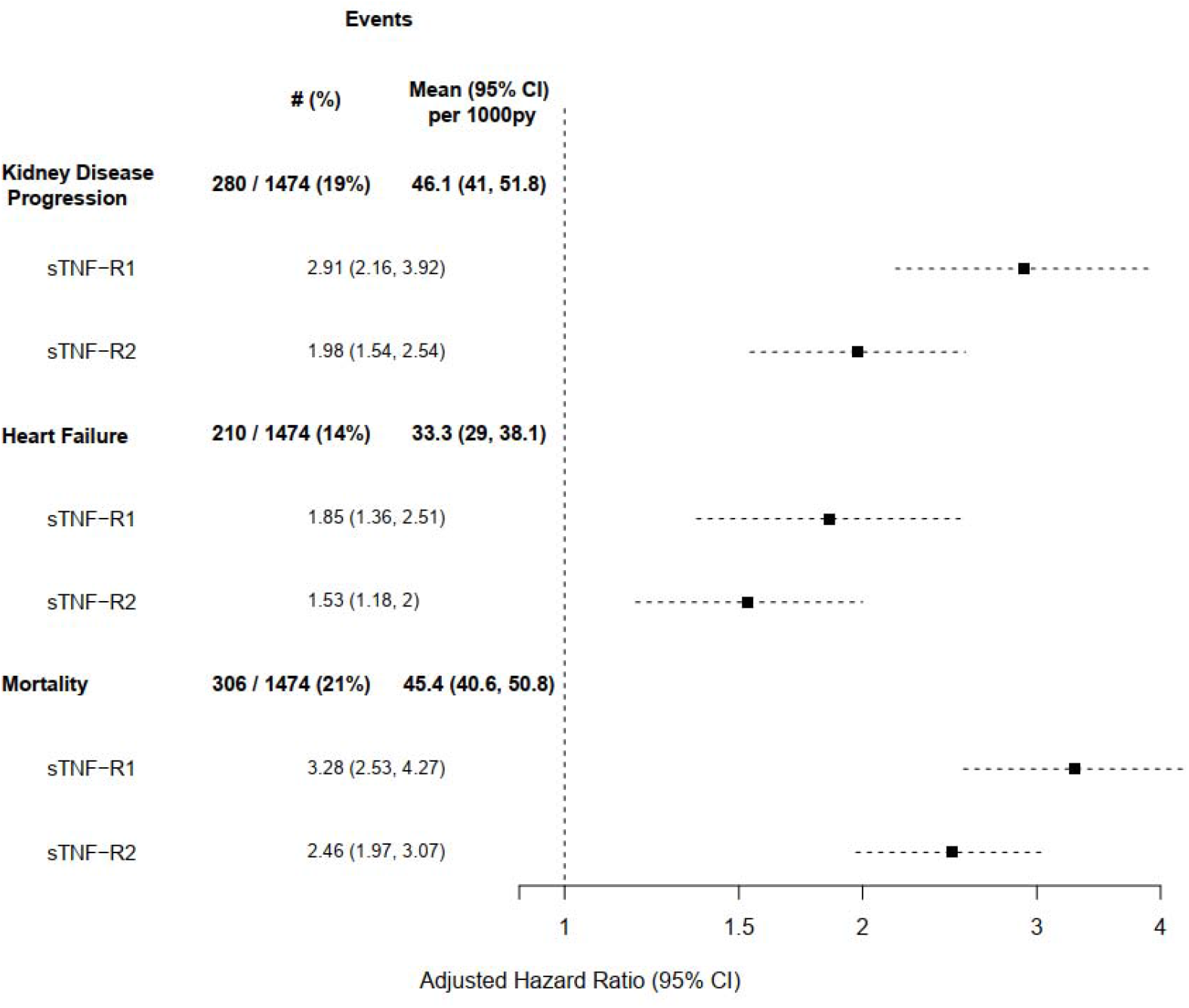
Forest plot demonstrating the adjusted HRs and 95% CIs for sTNFR1 and sTNFR2 at 3 months with adverse clinical outcomes in ASSESS-AKI. **Footnote:** Outcomes in a median follow-up of 4.4 (IQR 2.5, 5.7) years. Hazard ratios were adjusted for race, sex, age, BMI, smoking status, history COPD, CVD, AKI status (vs. no AKI), CKD status (vs. no CKD), sepsis during hospitalization, 3-month eGFR, 3-month UACR, 3-month CRP, and clinical site. Biomarkers were log-base 2 transformed.

**Figure 3.**
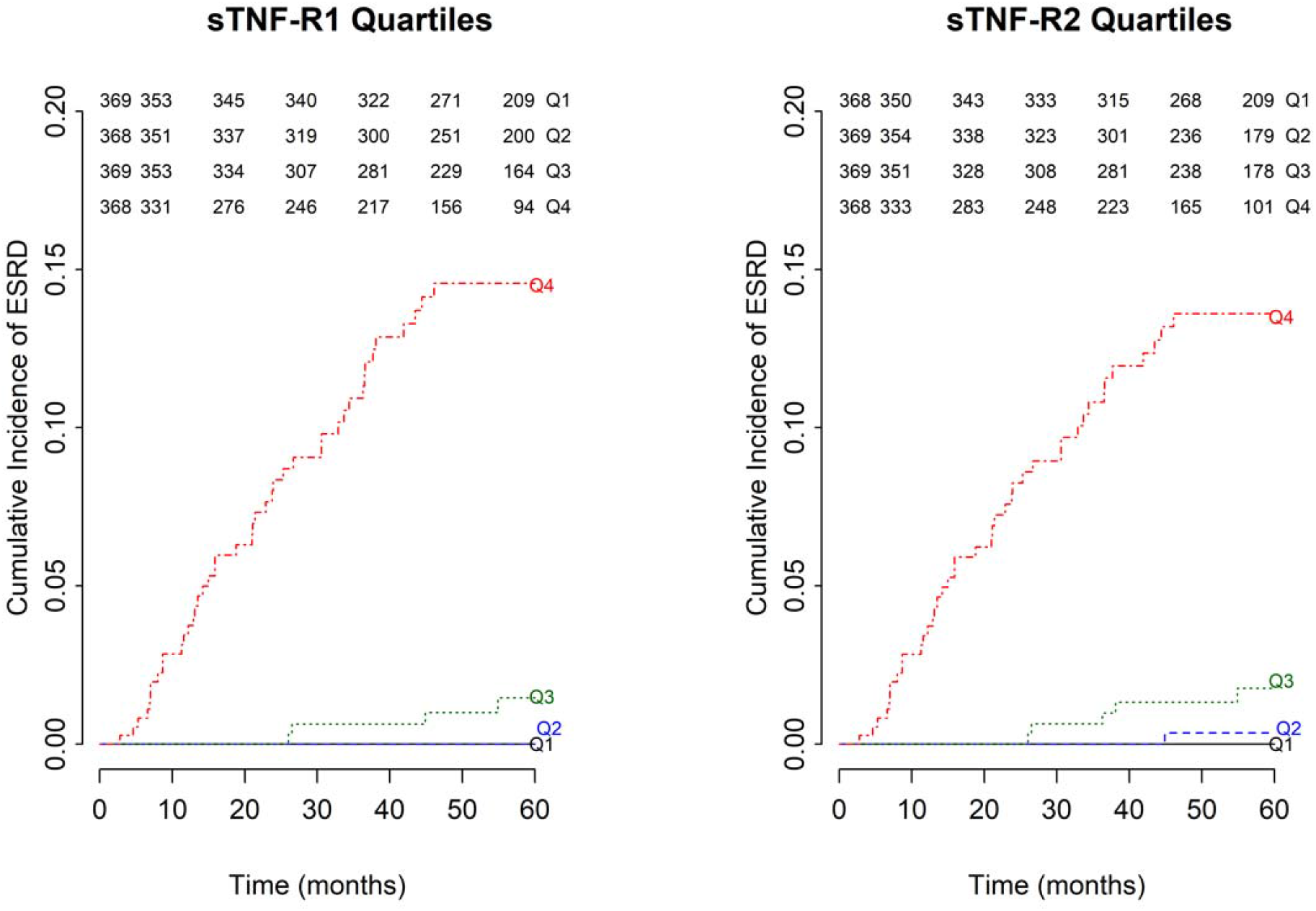
Time to ESKD events based upon sTNFR1 and sTNFR2 quartiles in ASSESS-AKI. **Footnote:** Median follow-up of 4.4 (IQR 2.5, 5.7) years.

### Association between sTNFR1 and sTNFR2 and heart failure in ASSESS-AKI

Among 1474 participants, 14% were hospitalized for heart failure during follow-up. Both sTNFR1 and sTNFR2 measured at 3 months of the index hospitalization were statistically and independently associated with heart failure hospitalization with aHR of 1.9 (95% CI: 1.4-2.5) and 1.5 (95% CI:1.2, 2) per doubling in sTNFR1 and sTNFR2, respectively, (**Table 2** and **Figure 2**) after adjusting for confounders.

### Association between plasma biomarkers and death in ASSESS-AKI

Overall, 21% of ASSESS-AKI participants died during follow-up, with an incidence of 45.4 (95% CI: 40.6-50.8) per 1000 person-years. Both biomarkers measured 3 months after the index hospitalization were independently associated with all-cause mortality with aHR 3.28 (95% CI: 2.5-4.2) per doubling for sTNFR1 and an aHR 2.5 (95% CI: 1.9, 3.1) per doubling for sTNFR2 after adjustment for confounders. Also, increasing risk of death was seen within higher biomarker quartiles (**Table 2 and Figure 2**).

### Plasma biomarkers in the prognosis of kidney disease progression, heart failure and death in the independent cohort: the ARID Study

ARID included 964 participants who were included in the secondary analysis (**Supplemental Figure 1**). This group was predominantly white (>90%) with mean age of 69 ± 11 years and a higher prevalence of CKD before hospitalization when compared to ASSESS-AKI (23.5% vs. 12.5%). Comparisons of baseline eGFR, clinical variables and biomarker cutoffs between both cohorts are described in **Supplemental Table 4** and **Supplemental Table 5**. Incident outcome rates in ARID and ASSESS-AKI are depicted in **Supplemental Figure 2**.

In ARID, 202 (21%) individuals developed the end point of kidney disease incidence or progression at 3 years compared to 175 (12%) participants in ASSESS-AKI. Participants had a 1.4-fold increased risk of the composite kidney endpoint per each doubling in the concentration of sTNFR1 (**Supplemental Figure 3**). There was no independent association between sTNFR2 and the kidney endpoint. In ARID, the strength of associations for sTNFR1 and sTNFR2 with heart failure and mortality were smaller in magnitude than those observed in ASSESS-AKI (**Supplemental Figure 3**).

## Discussion

Plasma levels of sTNFR1 and sTNFR2 measured 3 months after an acute hospitalization were independently associated with future progression of kidney disease. The strength of the association of sTNFR1 and sTNFR2 for all 3 outcomes were stronger in the US-based cohort (ASSESS-AKI) compared with the UK-based cohort (ARID). A rather novel finding was that these biomarkers were independently associated with heart failure in both cohorts, which highlights the potential importance of TNF-α signaling pathways in chronic heart and kidney disease. These biomarkers were also associated with death in ASSESS-AKI, which replicated for sTNFR2 in ARID. While the incidence of events was higher in those that had AKI during the index hospitalization, there was no evidence of effect modification by AKI on the association between sTNFRs and outcomes.

Prior studies have demonstrated both the severity and duration of AKI as well as the degree of post-AKI proteinuria strongly associate with future kidney events.^1,32,33,34^ We demonstrated that sTNFR1 and sTNFR2 measured 3 months after hospitalization independently associate with key long-term clinical kidney outcomes independently of kidney function (eGFR) and injury (UACR) parameters, as demonstrated in ASSESS-AKI. Furthermore, the association of sTNFRs with incident and progressive CKD has been demonstrated in diabetic kidney disease and CKD patients elsewhere.^13-15,17^ There was a marked gradation in risk for those in the 4^th^ vs 1^st^ quartiles of sTNFR1 and sTNFR2 for the development of kidney disease progression and ESKD. Our study adds to the evidence for the predictive role of sTNFRs after hospitalization and post-AKI outcomes, including recent data showing the utility of a multivariable model containing sTNFR1, sTNFR2, cystatin C and eGFR to discriminate those who have a significant worsening in kidney function in the three years following AKI.^16^ Also, sTNFRs were also strongly associated with outcomes in patients that did not experience AKI during the index hospitalization in ASSESS-AKI, raising the question of undetected ‘subclinical’ AKI or CKD that was not captured by creatinine criteria. This is important since subclinical, and milder forms of AKI may not be recognized by general practitioners.^10^ These relationships were qualitatively reproduced in ARID, comprised of 995 participants admitted to the general wards, with a higher frequency of baseline CKD compared to ASSESS-AKI (23.5% vs. 12.5%). Overall, the results for sTNFR1 and kidney outcomes seen in ARID were concordant to those obtained in ASSESS-AKI. While this independent verification is important to externally validate our results, different biomarker analytical platforms, sample size, follow-up period, and baseline CKD frequency could explain in part the heterogenous findings between cohorts.

We found that higher biomarker concentrations after discharge were associated with subsequent heart failure hospitalizations and mortality during the following 4.5 years, independent of other key confounders. The associations were stronger in ASSESS-AKI than in ARID. Serum TNFRs have been associated with adverse cardiovascular outcomes including myocardial infarction and heart failure in patients with coronary heart disease via complex cellular and molecular changes.^35,36,37^ Higher levels of these biomarkers have been associated with increased infarct size, left ventricular dysfunction and accelerated atherosclerosis. Therefore, these pathological changes can induce myocardial remodeling and contribute to development of heart failure.^37,38^ While higher sTNFR levels in CKD patients have been associated with the development of CVD,^39^ the relationship between these biomarkers and heart failure admissions after AKI is less known.

Overall, these findings may have direct application in post-hospitalization follow-up, especially when most AKI patients are seen by primary care physicians rather than nephrologists.^2,6^ Many patients, even those without ‘clinical’ kidney disease may have unappreciated renal parenchymal disease predisposing them to new episodes of AKI or have residual functional damage.^10^ Given the high discrimination for future kidney events (AUC 0.94) with either sTNFR1 or sTNFR2 and a small set of clinical variables, future studies could evaluate whether using sTNFR-enhanced risk assessment models could aid in triaging patients for nephrology consultation or for improved clinical care overall in those at highest risk for adverse kidney events. In this regard, measuring sTNFRs may be particularly helpful in for risk assessment in patients recovering from an acute illness who may have experienced loss of muscle mass, which may falsely bias eGFR higher post-illness.^40,41,42^

Study strengths include its large, multicenter, prospective design, with carefully matched participants, and broad inclusion criteria. The verification of some of the findings in an independent prospective cohort may favor the generalizability of the results. We collected data in a rigorous and structured manner and also systematically quantified post-AKI biomarkers at a uniform point in time relative to hospital discharge. In addition, we adjudicated clinical events after a long-term period of evaluation, which allows assessment for insidious conditions such as CKD. Nonetheless, study participants were predominantly white, high-risk individuals with prevalent comorbid conditions. In terms of limitations, it is unclear if these biomarkers could exhibit the same high performance in a less sick population. Further, the assays used in ASSESS and ARID were different, and thus, the biomarker concentration threshold at which the outcome risk is the highest could not be cross validated. Moreover, this study did not consider additional factors such as recurrent AKI or incidental hospitalizations which could arguably alter biomarker measurements and outcomes. Finally, the sample size of ARID was smaller than ASSESS-AKI.

In conclusion, sTNFR1 and sTNFR2 measured 3 months after hospital discharge were independently associated with kidney disease progression, heart failure and death, regardless of AKI status in the index admission. There was a linear association between incremental levels of these biomarkers and individual adverse outcomes. Evaluating post-discharge plasma sTNFR1 and sTNFR2 levels among hospital survivors, particularly in those who experienced AKI, may help in the risk-stratification of patients before the development of long-term kidney and cardiovascular sequelae.

## Supporting information

STROBE checklist

## Data Availability

All data produced in the present work are contained in the manuscript

https://repository.niddk.nih.gov/studies/assess-aki/

## Author contributions

*SGC and GVR contributed equally to the intellectual content and elaboration of this manuscript. SGC, GVR, HTP, and CRP drafted the manuscript. SGC, GVR, HTP, and CRP analyzed the data. For the ARID study team: Designed the study and oversaw its delivery: NMS; carried out data collection: RP; performed and oversaw biomarker analysis: REB, EM, CR; performed data analysis: MW; provided input on data analysis: NMS, REB, CR, MJK, EM. All authors participated in the conception and design of the study, acquisition or interpretation of the data, critical revision of the article for important intellectual content, and final approval of the version to be published.

## Acknowledgements

The authors would like to thank all of the ASSESS-AKI study participants, research coordinators, and support staff for making this study possible. The ASSESS-AKI was supported by cooperative agreements from the NIDDK (U01DK082223, U01DK082185, U01DK082192, and U01DK082183). We also acknowledge funding support from NIH grants R01HL085757, R01DK098233, R01DK101507, R01DK114014, K23DK100468, R03DK111881, R01DK093771, K01DK120783, P30DK079310, and P30DK114809. The ARID study was funded by a grant from Kidney Research UK (RP13/2015). The funder did not have any role in study design, data collection, analysis, reporting, or the decision to submit for publication. The ARID study would like to acknowledge the support provided by John Lamont and Peter FitzGerald from Randox Laboratories Ltd. The results opinions expressed in this paper do not necessarily reflect those of the National Institute of Diabetes Digestive and Kidney Diseases, the National Institutes of Health, the Department of health and Human Services or the government of the United States.

---

**Supplemental Table 1.** Patients’ characteristics on admission and by 3-month stratified by quartiles of plasma sTNFR2 concentrations in ASSESS-AKI.

|  | Plasma sTNFR2 Quartiles (pg/mL) |  |  |  | P- value |
| --- | --- | --- | --- | --- | --- |
| Characteristics on admission | Q1<br>2022, 4756<br>(N=368) | Q2<br>4758, 6651<br>(N=369) | Q3<br>6652, 9693<br>(N=369) | Q4<br>9710, 53522<br>(N=368) |  |
| Age (SD) | 60 (13.4) | 64.9 (12.5) | 66.9 (11.9) | 66.6 (11.8) | <.001 |
| Female | 129 (35%) | 124 (34%) | 146 (40%) | 139 (38%) | 0.3 |
| Black | 41 (11%) | 44 (12%) | 55 (15%) | 50 (14%) | 0.4 |
| Hispanic/Latino ethnicity | 8 (2%) | 10 (3%) | 11 (3%) | 6 (2%) | 0.6 |
| Smoker | 56 (15%) | 48 (13%) | 47 (13%) | 44 (12%) | 0.1 |
| Chronic liver disease | 2 (1%) | 8 (2%) | 18 (5%) | 29 (8%) | <.001 |
| Chronic obstructive pulmonary disease | 55 (15%) | 81 (22%) | 88 (24%) | 98 (27%) | 0.013 |
| Diabetes | 95 (26%) | 153 (41%) | 153 (41%) | 227 (62%) | <.001 |
| Sepsis | 25 (7%) | 40 (11%) | 30 (8%) | 45 (12%) | 0.04 |
| Heart failure | 34 (9%) | 61 (17%) | 92 (25%) | 125 (34%) | <.001 |
| Cardiovascular disease | 130 (35%) | 159 (43%) | 183 (50%) | 198 (54%) | <.001 |
| Chronic kidney disease | 34 (9%) | 100 (27%) | 175 (47%) | 275 (75%) | <.001 |
| Acute kidney injury | 131 (36%) | 159 (43%) | 192 (52%) | 244 (66%) | <.001 |
| 3-month follow-up measurements |  |  |  |  |  |
| Body mass index (kg/m<sup>2</sup>) | 29.9 (6.7) | 30.9 (7.5) | 31.1 (7.8) | 32.2 (8.35) | 0.007 |
| Estimated glomerular filtration rate, mL/min/1.73m<sup>2</sup> | 87.5 (18.3) | 75.9 (21.9) | 66.3 (21.7) | 47.9 (22.8) | <.001 |
| Urine albumin-to-creatinine ratio, mg/g (median, IQR) | 0.1 (0.1, 0.2) | 0.1 (0.1, 0.3) | 0.2 (0.1, 0.7) | 0.5 (0.1, 4.9) | <.001 |
| Plasma C reactive protein, mg/L (median, IQR) | 1.9 (0.9, 4.6) | 3.5 (1.7, 6.6) | 3.7 (1.8, 7.8) | 6.5 (3, 12.9) | <.001 |

**Supplemental Table 2.**
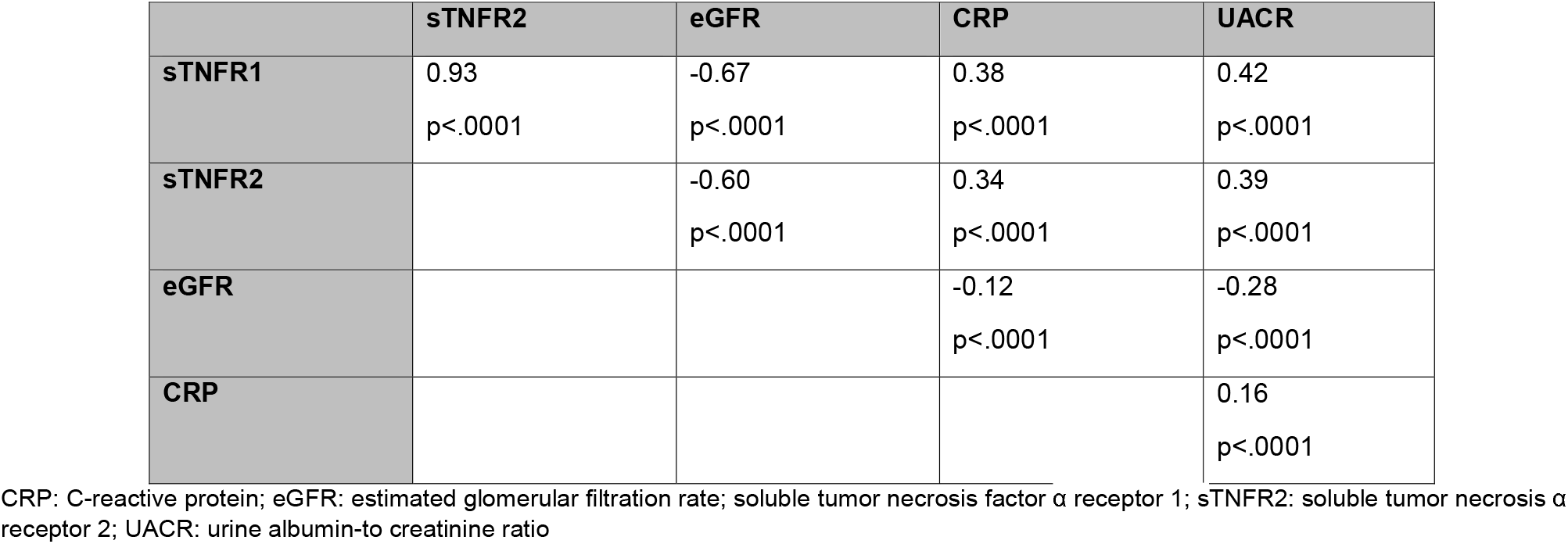
Spearman Correlation Table of sTNFR1 and sTNFR2 with eGFR, UACR and CRP in ASSESS-AKI.

**Supplemental Table 3.** Plasma sTNFR1 and sTNFR2 and kidney disease progression events stratified by AKI at the index hospitalization in ASSESS-AKI.

| Biomarker |  | Kidney Disease Progression Events, n (%) |  | Interaction p-value* |
| --- | --- | --- | --- | --- |
|  |  | AKI at Index Hospitalization |  |  |
|  |  | No (n=748) | Yes (n=726) |  |
| Overall |  | 89 (12%) | 191 (26%) | ... |
| TNFR1 (quartiles) | Q1 | 14 (6%) | 15 (12%) | 0.74 |
|  | Q2 | 28 (13%) | 36 (23%) |  |
|  | Q3 | 24 (14%) | 54 (28%) |  |
|  | Q4 | 23 (19%) | 86 (35%) |  |
| TNFR2 (quartiles) | Q1 | 17 (7%) | 16 (12%) | 0.54 |
|  | Q2 | 26 (12%) | 36 (23%) |  |
|  | Q3 | 26 (15%) | 55 (29%) |  |
|  | Q4 | 20 (16%) | 84 (34%) |  |

**Footnote:** Median follow up of 4.4 (IQR 2.5, 5.7) years. Patients that experienced AKI had higher concentrations of sTNFR1 [median (IQR) 3238 (2296, 4886)] and sTNFR2 [median (IQR) 7550 (5481, 11528)] at 3 months, compared to those without AKI: 2554 (1917, 3449) for sTNFR1, and 5835 (4424, 8137) for sTNFR2.
\*Interaction p-values are from the interaction term of a cox regression model of the biomarker, AKI at index hospitalization and the interaction between AKI and the biomarker.

**Supplemental Table 4.**
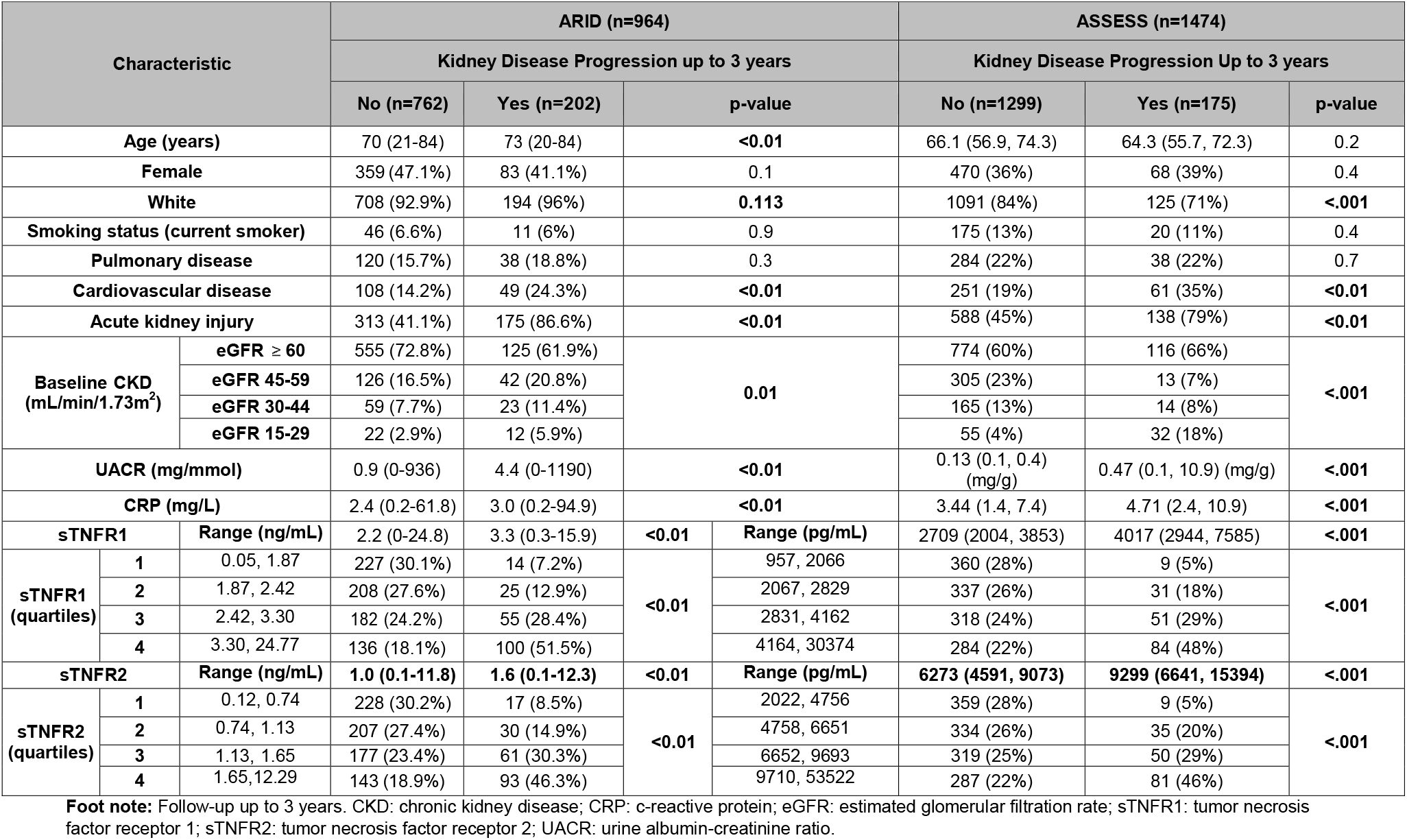
Patients’ characteristics by kidney disease progression up to 3 years in ARID vs. ASSESS-AKI.

**Supplemental Table 5.** Demographic and clinical characteristics by sTNFR1 and sTNFR2 quartiles in ARID.

| Characteristic |  | Plasma sTNFR1 (ng/mL) |  |  |  | P-value | Plasma sTNFR2 (ng/mL) |  |  |  | P-value |
| --- | --- | --- | --- | --- | --- | --- | --- | --- | --- | --- | --- |
|  |  | Q1<br>0.05, 1.87<br>(n=241) | Q2<br>1.87, 2.42<br>(n=233) | Q3<br>2.42, 3.30<br>(n=237) | Q4<br>3.30, 24.77<br>(n=236) |  | Q1<br>0.12, 0.74<br>(n=245) | Q2<br>0.74, 1.13<br>(n=237) | Q3<br>1.13, 1.65<br>(n=238) | Q4<br>1.65, 12.29<br>(n=236) |  |
| Age (years) |  | 66.0<br>(21.0-84.0) | 70.0<br>(20.0-84.0) | 74.0<br>(41.0-84.0) | 74.0<br>(42.0-84.0) |  | 68.0<br>(21.0-84.0) | 70.0<br>(33.0-84.0) | 73.0<br>(20.0-84.0) | 74.0<br>(42.0-84.0) | <0.001 |
| Female |  | 131 (54.4) | 106 (45.5) | 103 (43.5) | 96 (40.7) | 0.018 | 139 (56.7) | 107 (45.1) | 99 (41.6) | 91 (38.6) | <0.001 |
| White |  | 216 (89.6) | 216 (92.7) | 227 (95.8) | 226 (95.8) | 0.017 | 222 (90.6) | 223 (94.1) | 224 (94.1) | 225 (95.3) | 0.174 |
| Smoking status (current smoker) |  | 9 (3.8) | 23 (10.6) | 11 (5.1) | 13 (6.6) | 0.061 | 18 (7.7) | 13 (5.9) | 13 (5.9) | 13 (6.5) | 0.271 |
| Pulmonary disease |  | 34 (14.1) | 41 (17.6) | 39 (16.5) | 42 (17.8) | 0.684 | 28 (11.4) | 41 (17.3) | 47 (19.7) | 41 (17.4) | 0.084 |
| Cardiovascular disease |  | 19 (7.9) | 33 (14.2) | 35 (14.8) | 67 (28.4) | <0.001 | 18 (7.3) | 33 (13.9) | 38 (16.0) | 67 (28.4) | <0.001 |
| Acute kidney injury |  | 78 (32.4) | 101 (43.3) | 133 (56.1) | 163 (69.1) | <0.001 | 77 (31.4) | 106 (44.7) | 143 (60.1) | 158 (66.9) | <0.001 |
| Baseline CKD stage (mL/min) | eGFR ≥ 60 | 228 (94.6) | 190 (81.5) | 157 (66.2) | 101 (42.8) | <0.001 | 224 (91.4) | 186 (78.5) | 159 (66.8) | 107 (45.3) | <0.001 |
|  | eGFR 45-59 | 12 (5.0) | 37 (15.9) | 60 (25.3) | 57 (24.2) |  | 17 (6.9) | 43 (18.1) | 56 (23.5) | 50 (21.2) |  |
|  | eGFR 30-44 | 1 (0.4) | 5 (2.1) | 19 (8.0) | 50 (21.2) |  | 4 (1.6) | 8 (3.4) | 19 (8.0) | 49 (20.8) |  |
|  | eGFR 15-29 | 0 (0.0) | 1 (0.4) | 1 (0.4) | 28 (11.9) |  | 0 (0.0) | 0 (0.0) | 4 (1.7) | 30 (12.7) |  |
| UACR (mg/mmol) |  | 0.7<br>(0.0-101) | 0.9<br>(0.0-101) | 1.4<br>(0.0-494) | 5.8<br>(0.0-1190) | <0.001 | 0.7<br>(0.0-494) | 0.8<br>(0.0-142) | 1.4<br>(0.0-196) | 5.3<br>(0.0-1190) | <0.001 |
| CRP (mg/L) |  | 1.6<br>(0.2-31.3) | 1.9<br>(0.2-56.4) | 2.9<br>(0.5-57.4) | 4.7<br>(0.4-94.9) | <0.001 | 1.6<br>(0.2-56.4) | 2.3<br>(0.2-45.5) | 2.6<br>(0.4-57.4) | 4.5<br>(0.5-94.9) | <0.001 |

**Foot note:** Follow-up up to 3 years. CKD: chronic kidney disease; CRP: c-reactive protein; eGFR: estimated glomerular filtration rate; sTNFR1: tumor necrosis factor receptor 1; sTNFR2: tumor necrosis factor receptor 2; UACR: urine albumin-creatinine ratio.

**Supplemental Table 6.**
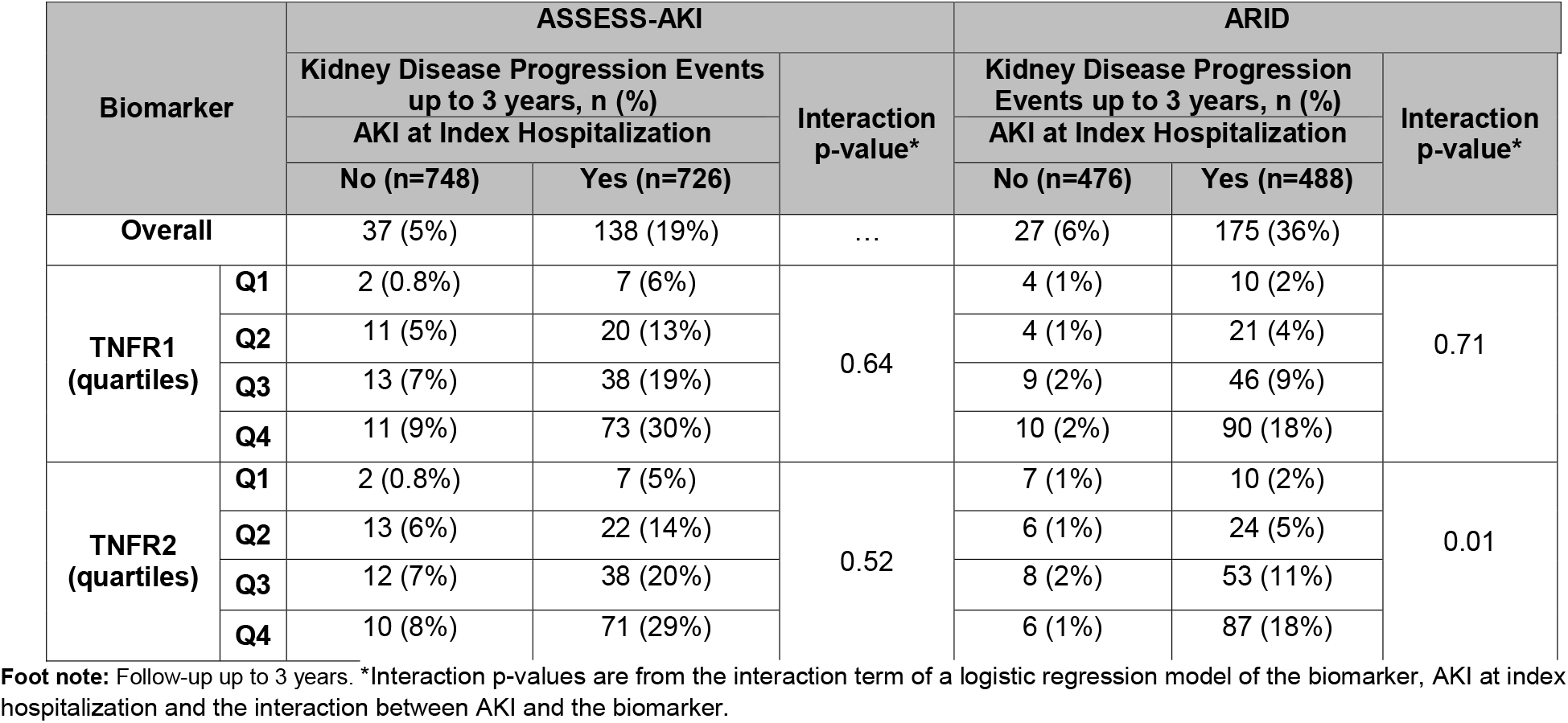
sTNFR1 and sTNFR2 and kidney disease progression events up to 3 years stratified by AKI at Index Hospitalization in ASSESS-AKI and ARID.

**Supplementary Table 7.** Comparison of patients included and excluded in ASSESS-AKI analysis.

|  | **Excluded**<br>**(N=64)** | **Included**<br>**(N=1474)** | **P\*\*** |
| --- | --- | --- | --- |
| **Characteristics on admission** |  |  |  |
| **Age** | 63.89 (13.36) | 64.58 (12.69) | 0.587 |
| **Female** | 36 (56%) | 538 (36%) | **0.001** |
| **Black** | 14 (22%) | 190 (13%) | **0.038** |
| **Hispanic/Latino Ethnicity** | 3 (5%) | 35 (2%) | 0.243 |
| **Smoker** | 7 (11%) | 195 (13%) | 0.856 |
| **Chronic liver disease** | 3 (5%) | 57 (4%) | 0.926 |
| **Chronic obstructive pulmonary disease** | 13 (20%) | 322 (22%) | 0.201 |
| **Diabetes** | 30 (47%) | 628 (43%) | 0.499 |
| **Sepsis** | 4 (6%) | 140 (9%) | 0.383 |
| **Heart failure** | 15 (23%) | 312 (21%) | 0.891 |
| **Cardiovascular disease** | 23 (36%) | 670 (45%) | 0.298 |
| **Chronic Kidney Disease** | 28 (44%) | 584 (40%) | 0.509 |
| **Acute Kidney Injury** | 43 (67%) | 726 (49%) | 0.005 |
| **3 month follow-up measurements** |  |  |  |
| **Body mass index (kg/m<sup>2</sup>)** | 31.78 (9.39)<br>N=64 | 31.02 (7.63)<br>N=1472 | 0.824 |
| **Estimated glomerular filtration rate, mL/min/1.73m<sup>2</sup>** | 63.22 (27.92)<br>N=64 | 69.43 (25.7)<br>N=1474 | 0.076 |
| **Urine albumin-to-creatinine ratio, mg/g (median, IQR)** | 0.32 (0.1, 1.23)<br>N=64 | 0.14 (0.07, 0.59)<br>N=1467 | **0.005** |
| **Plasma C reactive protein, mg/L (median, IQR)** | 3.07 (1.81, 10.73)<br>N=21 | 3.56 (1.5, 7.61)<br>N=1463 | 0.995 |

**Supplementary Figure 1.**
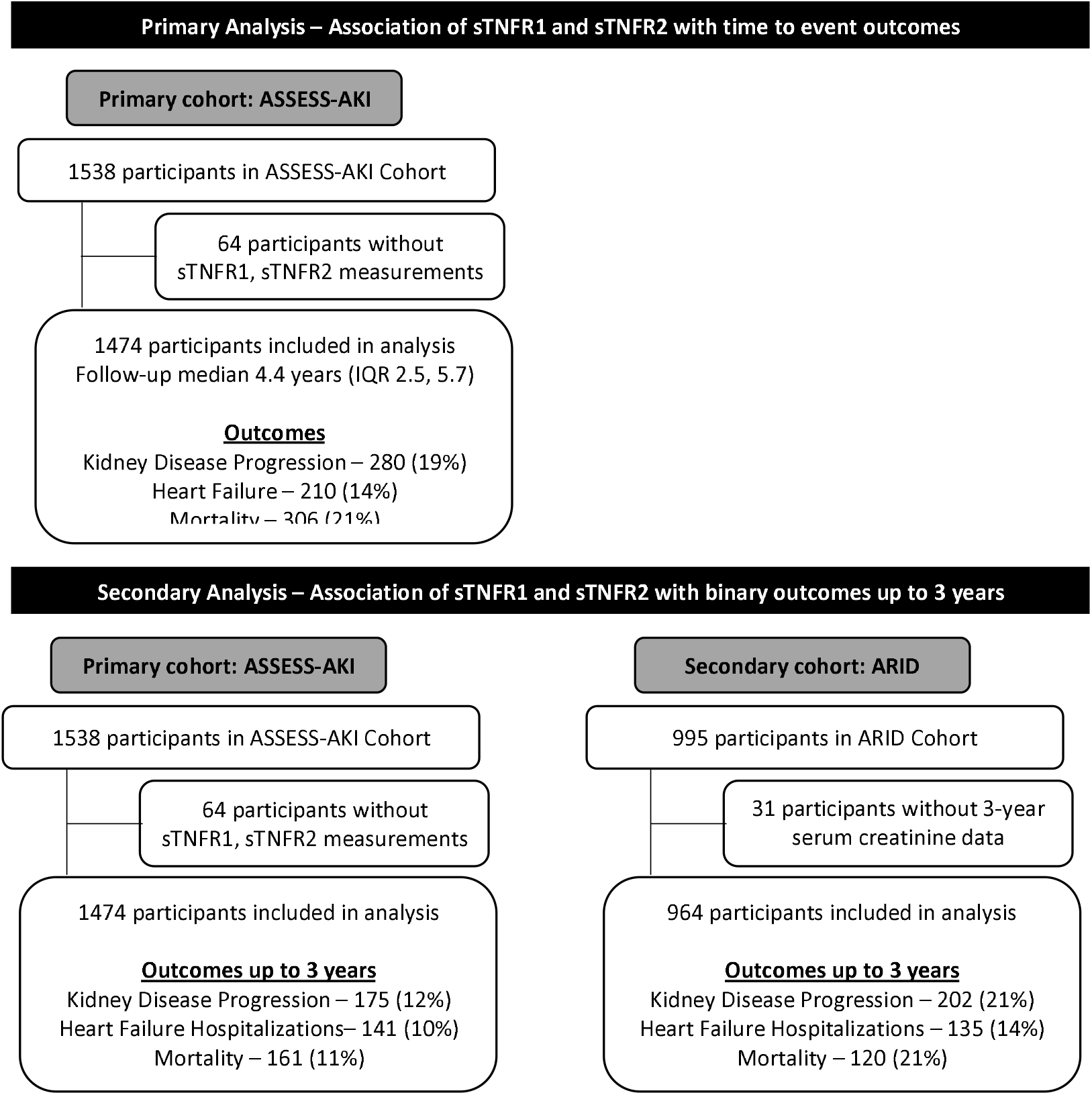
Flow diagram for ASSESS-AKI and ARID

**Supplementary Figure 2.**
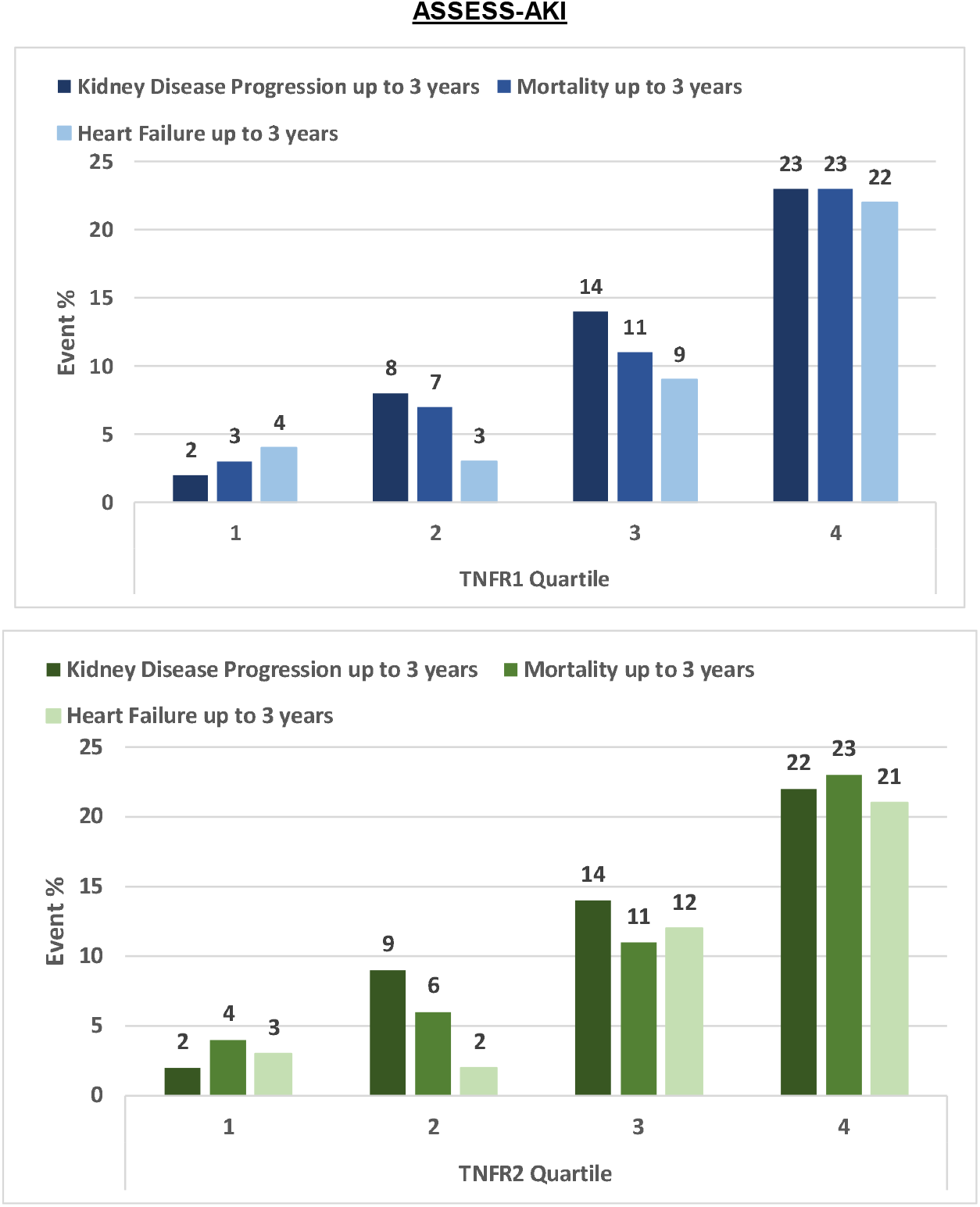
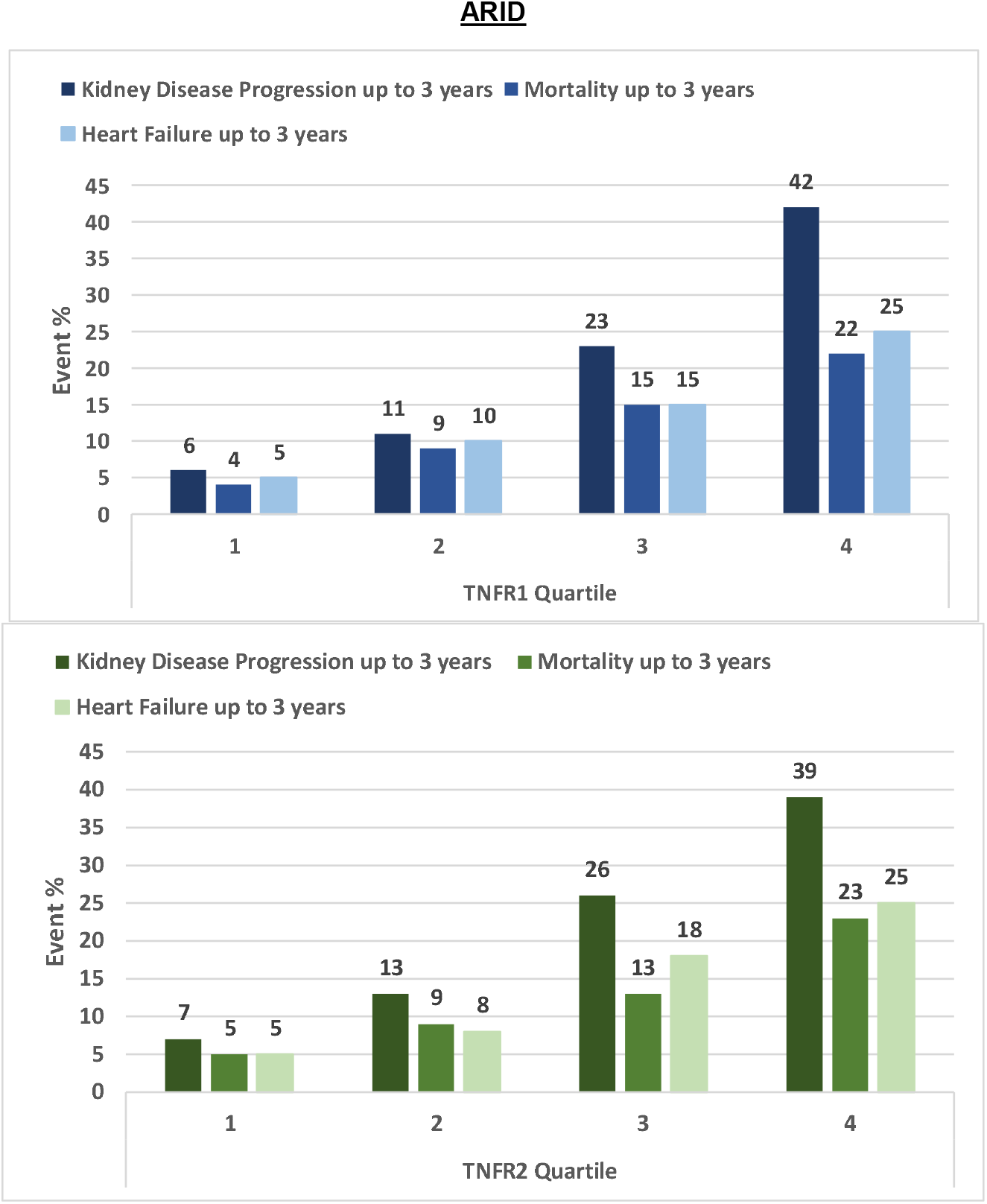
Event rates of 3-year outcomes by quartiles of plasma sTNFR1 and sTNFR2 concentrations in ASSESS-AKI and ARID.

**Supplementary Figure 3.**
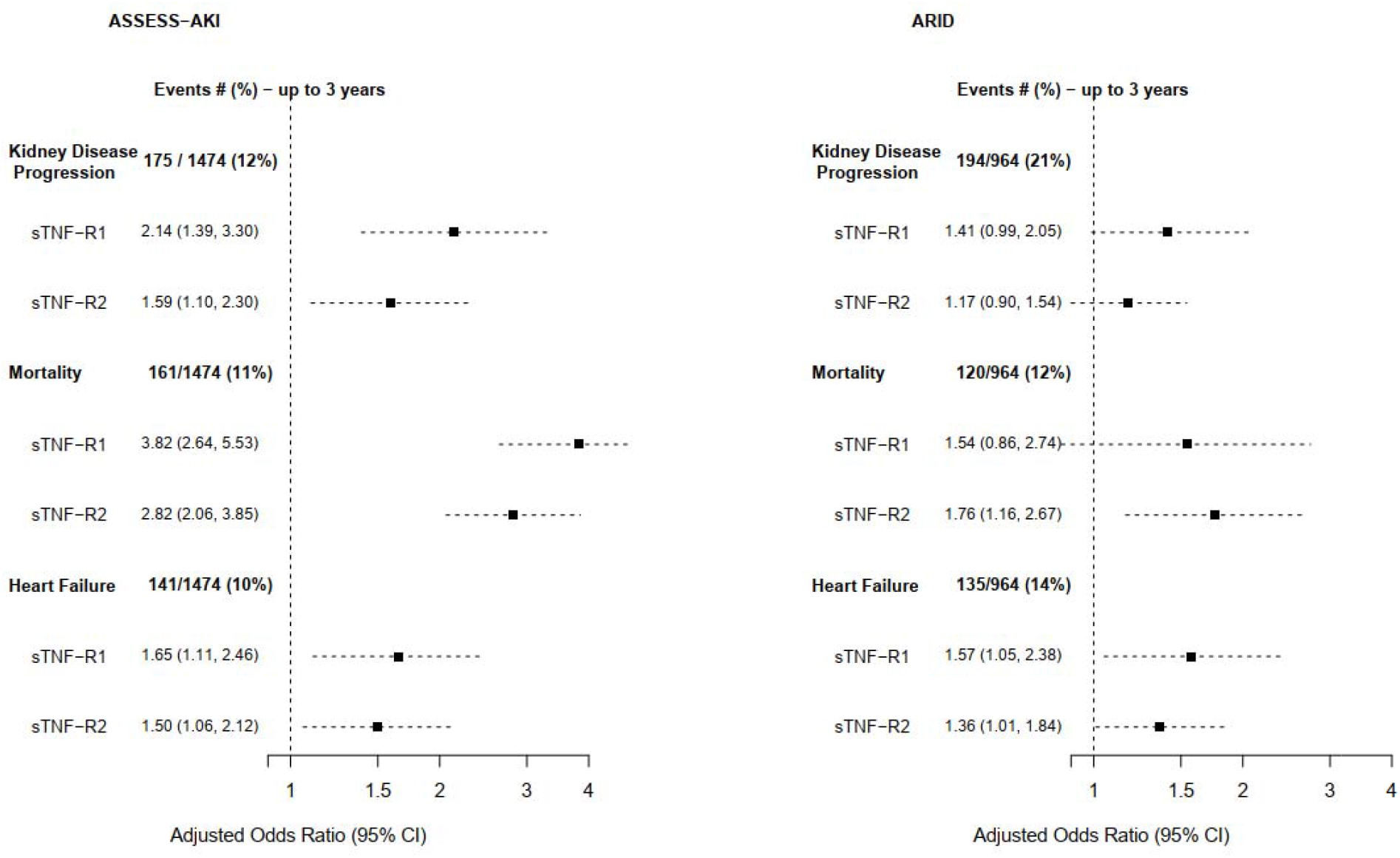
Forest plot demonstrating the adjusted ORs and 95% CIs for sTNFR1 and sTNFR2 measured at 3 months with adverse clinical outcomes in ASSESS-AKI and ARID. **Foot note:** Odds ratios were adjusted for white race (yes/no), sex, age, smoking status, history of COPD, CRP, baseline CKD status (yes/no), AKI status, 3-month GFR, 3-month UACR

