## Supplementary material for "Plasma Soluble Tumor Necrosis Factor Receptor Concentrations and Clinical Events after Hospitalization: Findings from ASSESS-AKI and ARID studies": STROBE checklist

STROBE Statement—Checklist of items that should be included in reports of ***cohort studies***

|  | Item No | Recommendation |
| --- | --- | --- |
| **Title and abstract** | 1 | (*a*) Indicate the study’s design with a commonly used term in the title or the abstract  Our title is: ‘Plasma Soluble Tumor Necrosis Factor Receptor Concentrations and Clinical Events after Hospitalization: Findings from ASSESS-AKI and ARID studies’ – ASSESS-AKI and ARID are two well known prospective cohorts that evaluate AKI outcomes after hospitalization. |
|  |  | (*b*) Provide in the abstract an informative and balanced summary of what was done and what was found – A comprehensive summary was provided. |
| Introduction | | |
| Background/rationale | 2 | Explain the scientific background and rationale for the investigation being reported  The first two paragraphs set what’s the current knowledge of our problem and what are the remaining challenges. The next paragraphs present what’s our study project about and how we plan to fill the gap in knowledge. |
| Objectives | 3 | State specific objectives, including any prespecified hypotheses  We included ‘We sought to further explore the association between plasma sTNFR1 and sTNFR2 concentrations post-discharge with long-term kidney-related outcomes, heart failure hospitalizations, and mortality in ASSESS-AKI and ARID among patients who did and did not have AKI. Furthermore, we aimed to assess whether measuring sTNFR1 and sTNFR2 could add to the performance of clinical models.’ |
| Methods | | |
| Study design | 4 | Present key elements of study design early in the paper  Prospective, matched, parallel cohort study including AKI survivors who were followed-up in a protocolized fashion. The association between 3-month biomarkers and longitudinal outcomes was assessed. Also, we have a validation cohort that was incorporated to see if the findings from the primary cohort replicated. |
| Setting | 5 | Describe the setting, locations, and relevant dates, including periods of recruitment, exposure, follow-up, and data collection  The primary cohort (ASSESS-AKI): US-based, multicentric, patients enrolled from December 2009 and February 2015. Mean follow-up period: 4.4 years  Replication cohort (ARID): UK-based, patients enrolled from September 2011 and October 2012. Mean follow-up: 3 years. |
| Participants | 6 | (*a*) Give the eligibility criteria, and the sources and methods of selection of participants. Describe methods of follow-up  Participants were age ≥18 years and had pre-admission serum creatinine levels that were obtained within 1 year of the index hospitalization in the outpatient setting or non-emergency department visit. Detailed inclusion criteria has been described elsewhere. Patients had to survive to complete an in-person visit at 3 months after discharge. |
|  |  | (*b*) For matched studies, give matching criteria and number of exposed and unexposed  Participants with AKI were matched at each participating institution based upon baseline characteristics, CKD, AKI stage, and urine studies. |
| Variables | 7 | Clearly define all outcomes, exposures, predictors, potential confounders, and effect modifiers. Give diagnostic criteria, if applicable.  Outcomes: death, kidney disease incidence and progression (incident CKD, progression of CKD, development of ESRD or transplant), heart failure hospitalizations and all-cause mortality.  Exposure: biomarkers (TNFR1, TNFR2)  Confounders: age, BMI, diabetes mellitus status, AKI stage, baseline CKD (eGFR, albuminuria), sepsis, baseline CVD, others. All these were adjusted during regression models.  Effect modifiers: we evaluated whether AKI interacted with the association between biomarkers and outcomes. |
| Data sources/ measurement | 8* | For each variable of interest, give sources of data and details of methods of assessment (measurement). Describe comparability of assessment methods if there is more than one group  Variables were obtained from chart review. Biomarkers in the primary cohort were measured via Meso Scale Discovery Platform and in ARID, we used Randox’s multiplexed Bioship Arrays. Both are well validated methods for detecting biomarkers. |
| Bias | 9 | Describe any efforts to address potential sources of bias  Selection bias: we tried to incorporate AKI patients from different institutions and to validate our findings with a European cohort to improve the generalizability of our findings. Both cohorts were protocolized, and similar in design. Number of participants who were lost to follow-up is minimal, limiting attrition bias. |
| Study size | 10 | Explain how the study size was arrived at 1474. Power calculations for alpha: 0.05 and power of 80% |
| Quantitative variables | 11 | Explain how quantitative variables were handled in the analyses. If applicable, describe which groupings were chosen and why  Quantitative variables such as biomarkers were evaluated as continuous (long2-transformation) and by quartiles |
| Statistical methods | 12 | (*a*) Describe all statistical methods, including those used to control for confounding  ‘Descriptive statistics for continuous variables were reported as mean (standard deviation) or median [interquartile range (IQR)] when appropriate. Categorical variables were presented as frequencies. Clinical and demographic differences were evaluated through the Kruskal-Wallis test or Chi-square tests. The primary analysis consisted in the evaluation of clinical outcomes including kidney disease progression, heart failure hospitalizations, and mortality in ASSESS-AKI. We used Cox proportional hazard models that were adjusted for race, sex, age, body mass index (BMI), smoking status, history of chronic obstructive pulmonary disease (COPD), cardiovascular disease (CVD), AKI status (vs. no AKI), CKD status (vs. no CKD), sepsis, 3-month eGFR, 3-month urine albumin creatinine ratio (UACR), 3-month c-reactive protein (CRP), and diabetes mellitus status. Hazard ratios (HR) and 95% confidence intervals (CI) multivariable models are reported. In addition, we assessed the receiver operating characteristic (ROC) curves of sTNFR1 and sTNFR2 in ASSESS-AKI for the discrimination of kidney events alone and when added to a clinical model comprised of eGFR, UACR, and C-reactive protein at 3 months, AKI at the index hospitalization, race/ethnicity, age, smoking, and COPD.’ |
|  |  | (*b*) Describe any methods used to examine subgroups and interactions  We evaluated the interaction of AKI (AKI yes/no) on the association between biomarkers and outcomes. |
|  |  | (*c*) Explain how missing data were addressed: Missing data was minimal. Description of such group of patients was incorporated in the manuscript. |
|  |  | (*d*) If applicable, explain how loss to follow-up was addressed. Those patients were excluded from the final analysis. |
|  |  | (*e*) Describe any sensitivity analyses. Sensitivity analyses were not conducted. |
| Results | | |
| Participants | 13* | (a) Report numbers of individuals at each stage of study—eg numbers potentially eligible, examined for eligibility, confirmed eligible, included in the study, completing follow-up, and analysed – Included in the patient flow-chart |
|  |  | (b) Give reasons for non-participation at each stage – included and described in detail |
|  |  | (c) Consider use of a flow diagram – included in the manuscript |
| Descriptive data | 14* | (a) Give characteristics of study participants (eg demographic, clinical, social) and information on exposures and potential confounders – included in the manuscript |
|  |  | (b) Indicate number of participants with missing data for each variable of interest  – included in the manuscript |
|  |  | (c) Summarise follow-up time (eg, average and total amount)  – included in the manuscript |
| Outcome data | 15* | Report numbers of outcome events or summary measures over time  – included in the manuscript, described above |
| Main results | 16 | (*a*) Give unadjusted estimates and, if applicable, confounder-adjusted estimates and their precision (eg, 95% confidence interval). Make clear which confounders were adjusted for and why they were included  – included in the manuscript, described in statistical methods. |
|  |  | (*b*) Report category boundaries when continuous variables were categorized  – included in the manuscript. See supplemental tables. |
|  |  | (*c*) If relevant, consider translating estimates of relative risk into absolute risk for a meaningful time period – not applicable |
| Other analyses | 17 | Report other analyses done—eg analyses of subgroups and interactions, and sensitivity analyses – interaction analyses included in the manuscript and described in detail |
| Discussion | | |
| Key results | 18 | Summarise key results with reference to study objectives – included in the manuscript. Biomarkers were independently associated with each of the three outcomes. |
| Limitations | 19 | Discuss limitations of the study, taking into account sources of potential bias or imprecision. Discuss both direction and magnitude of any potential bias  – included in the manuscript (discussion section) |
| Interpretation | 20 | Give a cautious overall interpretation of results considering objectives, limitations, multiplicity of analyses, results from similar studies, and other relevant evidence  – included in the manuscript |
| Generalisability | 21 | Discuss the generalisability (external validity) of the study results  – included in the manuscript |
| Other information | | |
| Funding | 22 | Give the source of funding and the role of the funders for the present study and, if applicable, for the original study on which the present article is based- A careful description of the author funding was included within the manuscript in acknowledgements. |

*Give information separately for exposed and unexposed groups.

**Note:** An Explanation and Elaboration article discusses each checklist item and gives methodological background and published examples of transparent reporting. The STROBE checklist is best used in conjunction with this article (freely available on the Web sites of PLoS Medicine at http://www.plosmedicine.org/, Annals of Internal Medicine at http://www.annals.org/, and Epidemiology at http://www.epidem.com/). Information on the STROBE Initiative is available at http://www.strobe-statement.org.
